# Metastasis Extraction from NSCLC Clinical Notes: A Retrospective Comparative Evaluation of Large Language Model-Based Classification

**DOI:** 10.64898/2026.04.27.26351872

**Authors:** Sweta Balaji, Kiersten Campbell, Rou-Zhen Chen, Daniel Smith, Matthew A. Reyna, Abeed Sarker, Joshua D. Wallach, Ravi B. Parikh, Selen Bozkurt

**Affiliations:** Department of Biomedical Informatics, Emory University, Atlanta, GA, USA; Department of Epidemiology, Rollins School of Public Health, Emory University, Atlanta, GA, USA; Winship Cancer Institute, Emory University, Atlanta, GA, USA; Department of Hematology and Medical Oncology, School of Medicine, Emory University, Atlanta, GA, USA

## Abstract

**Background:** Identification of metastasis status in non-small cell lung cancer (NSCLC) is a critical part of understanding disease prognosis, treatment courses, trial eligibility, and population-level cancer surveillance. However, metastasis record are inconsistently recorded in structured cancer registry fields, since manual abstraction of clinical notes is often a resource intensive and error-prone process. This challenge highlights an opportunity for leveraging large language models (LLMs) to conduct high-scale metastasis extraction from real-world clinical documentation.

**Objective:** We conducted a retrospective, multi-cohort comparative evaluation of three distinct LLMs for two independent classification tasks: overall metastasis presence at any site and brain/CNS metastasis presence. We evaluated model performance on two independent NSCLC cohorts: (1) a registry-linked cohort used for model development and validation and (2) an independent cohort with manual note-level annotations for additional validation. We further explored whether our methods could analyze clinical documentation and recover missing or outdated metastasis information in structured registry labels.

**Methods:** Patient cohorts were derived from the Winship Cancer Institute. Cohort 1 (n=579 patients; 24,887 notes across 69 note types; 2023-2025) used registry-linked metastasis fields as the reference standard. Cohort 2 (n=22 patients; 644 radiology notes; 2010-2021) was drawn from two completed randomized trials and used dual-annotator manual labels (Cohen’s &[kappa]: 0.93 overall metastasis, 0.88 CNS metastasis) as the reference standard. We fine-tuned the GatorTron-base encoder model for each independent binary classification task, respectively. We evaluated MedGemma-27B-text and Llama 3.1-70B using zero-shot prompting. A separate cohort of 675 patients with missing or unknown registry labels was used for an exploratory missingness-recovery analysis, validated against manual annotations of a random subsample.

**Results:** More than half (54%) of initially identified Cohort 1 patients had missing or unknown registry metastasis labels. For overall metastasis, fine-tuned MedGemma demonstrated the best performance in overall metastasis classification (Cohort 1: F1=0.80, Cohort 2 patient level: F1=1.0, Cohort 2 note level: F1=0.93). For brain/CNS metastasis, Llama3 performed best in both cohorts (Cohort 1: F1=0.79, Cohort 2 patient-level: F1=0.93, Cohort 2 note-level: F1=0.86). The fine-tuned GatorTron model showed strong performance for classification of overall metastasis in Cohort 1 (F1=0.72). Error analysis indicated that most model errors reflected incomplete registry labels, ambiguous clinical language, or missing documentation rather than true model errors. In the exploratory recovery analysis, model predictions agreed with manual annotations at accuracy=0.90 and F1=0.89.

**Conclusions:** All models demonstrated relatively high performance. The zero-shot generative models were more robust to nuanced documentation and context-dependent brain/CNS metastasis extraction. The fine-tuned encoder model demonstrated strong classification performance but may have been limited by potential inaccuracies in the registry reference standards during model training. This study further demonstrated the potential of LLMs in recovering clinically plausible structured labels from narrative text, complementing cancer registries for metastasis ascertainment.

## Introduction

Non-small cell lung cancer (NSCLC) is the leading cause of cancer-related mortality worldwide, with prognosis and treatment shaped by the presence and extent of metastatic disease at diagnosis and throughout the clinical course [1–3]. Among the most consequential and clinically complex sites of spread is the central nervous system (CNS) [4]. Brain metastasis develops in up to 50% of NSCLC patients over the course of their disease, carrying distinct prognostic implications, driving treatment decisions, and serving as a key eligibility criterion in oncology trials [5,6]. Accurate ascertainment of metastatic status is crucial for determining eligibility for systemic therapies, radiation planning, enrollment in clinical trials, and cancer surveillance [7,8]. These realities underscore that metastasis information, particularly CNS metastasis, must be captured accurately and comprehensively.

However, metastatic status is inconsistently recorded in structured electronic health record (EHR) fields and is often dispersed across radiology reports, oncology progress notes, discharge documentation, and other free-text clinical narratives [9]. Cancer registries, the primary infrastructure for population-level cancer surveillance, rely on manual review by trained cancer registrars, an approach that is resource-intensive, subject to human error, and limited in its ability to capture disease progression documented after the initial abstraction window [10]. These gaps in data capture can have meaningful consequences. In a large National Cancer Database analysis of more than 1.2 million patients with NSCLC, 71.0% had missing data in at least one variable of interest, including cancer stage and treatment [11]. Missing data were especially prevalent among patients with lower 2-year overall survival, indicating that incomplete documentation may have been more prevalent for patients with severe illnesses or complex, metastatic cases [11]. Thus, studies utilizing cancer registry data may be biased if missingness is ignored [11]. Another limitation of cancer registries is that data often reflect disease status primarily at the time of initial diagnosis and do not capture metastatic progression documented later in the clinical course [12,13]. In contrast, clinical notes provide longitudinally detailed records of disease evolution, including imaging interpretation, treatment modifications, and clinician assessments across the patient journey. Manual abstraction of clinical notes is a lengthy, resource-intensive process that is prone to human error and may not comprehensively capture disease complexities documented throughout the course of a patient’s medical history [14]. These challenges highlight an opportunity for informatics approaches, namely natural language processing (NLP) methods, to enable large scale extraction of metastasis records from real-world clinical documentation, improving accuracy and comprehensiveness of existing structured data sources [15].

There has been rapid evolution of NLP methods in the field of clinical information extraction. Early rule-based methods leveraged medical ontologies to create structured rules for detection of relevant information in unstructured text [16]. This method continues to be used today, particularly for text filtering and standardization [17]. Rule-based methods have previously demonstrated high precision and interpretability but with limited ease of implementation and generalizability to notes beyond the study context [18,19]. Its limited generalizability is characterized by its dependence on specific linguistic patterns, along with requiring specialized human expertise to create custom rules [20]. These rules can be affected by unique documentation style and phrasing of the data used [20]. Machine learning (ML) approaches demonstrated improvement upon rule-based systems due to their ability to automatically learn linguistic patterns and complexities rather than solely capturing patterns based on predefined rules [21]. Transformer-based ML models, specifically, have demonstrated strong performance in real-world data and can be fine-tuned for use in various clinical contexts [21]. Transformers pretrained on clinical and medical data have further demonstrated immense promise in detecting semantic interactions and complex, varied phrasing in clinical text [22,23]. These models, further optimized on smaller clinical cohorts, have shown to outperform traditional NLP and ML approaches, including regular expression-based text searches and classic supervised learning approaches [24]. However, lower performance has been observed when applying these models to heterogeneous note types or switching institutions from which the notes are sourced. Furthermore, differences in vocabulary and documentation style can be seen to lower performance in these transformer models [23]. More recently, generative large language models (LLMs), including both domain-adapted and general-purpose instruction-tuned variants, have demonstrated strong performance on a range of clinical extraction tasks [11,25–27]. However, since these models generate free text, their outputs can be inconsistent and hard to generalize for clinical extraction tasks compared to BERT models [28]. These models also require a substantial amount of memory, which can detract from widespread use in clinical settings [23].

Despite this progress, critical gaps remain, particularly in the types of clinical text analyzed for extraction tasks. First, a recent systematic review of LLM applications in clinical medicine found that over three-quarters of included studies did not conduct analyses on real EHR data, relying instead on simulated clinical scenarios or synthetic notes [29]. This limits generalizability of findings to the nuanced documentation encountered in routine oncology practice. Second, few studies have directly compared fine-tuned domain-adapted encoder models, for supervised classification, against instruction-tuned generative models, particularly for information extraction from real-world oncology notes [23,28]. This raises the question of which method is more effective. Third, and particularly for NSCLC, brain and CNS metastasis extraction is underexamined as a distinct NLP task, despite its clinical importance. Given the complex language and the plethora of terminologies used to document different sites of CNS metastasis, this extraction task further tests the ability of NLP methods or LLMs in detecting and distinguishing between nuanced phrases and site-specific verbiage.

To address these gaps, we conducted a retrospective multi-cohort comparative evaluation of LLM-based methods for extracting overall metastasis and brain/CNS metastasis from real-world NSCLC clinical notes. Our primary aim was to evaluate model performance on a registry-linked reference standard in an institutional cohort and further validate findings in an independent cohort from an earlier documentation period with manual note-level annotations. We further sought to explore whether our methods could analyze clinical documentation and recover metastasis information missing from structured registry fields. Ultimately, our goal is to support the design of scalable NLP pipelines capable of supporting cancer registry abstraction, improving metastasis ascertainment in EHR-derived datasets, and enabling more complete and accurate identification of NSCLC patients in clinical trial eligibility screening.

## Methods

### A. Study Design and Setting

We conducted a retrospective comparative evaluation of three LLM-based methods for extracting (1) overall metastasis at any site and (2) brain/CNS metastasis from real-world clinical notes in NSCLC patients. We leveraged two cohorts with distinct but complementary roles in model development and validation, in addition to a dataset with missing metastasis records to explore the LLM’s capabilities for data recovery. All analyses were conducted under a waived-consent protocol approved by the Emory University Institutional Review Board.

### B. Data Sources and Cohorts

#### Cohort 1: Primary Registry-Linked Development Cohort

Cohort 1 was used for model development and internal validation. It was derived from the Winship Cancer Registry, linked to the Georgia Comprehensive Cancer Registry. Inclusion was restricted to patients for whom Winship Cancer Institute was the institution of record for diagnosis and/or first-course treatment, with both registry data and linked clinical notes available from 2023 through 2025.

#### Cohort construction and exclusions

Initial extraction yielded 1,260 NSCLC patients with 62,191 notes across 81 note types and 12,174 encounters. Patients were excluded from model development if at least one registry metastasis field was missing (*n* = 663) or labeled unknown with no confirmed positive site (*n* = 12), yielding 585 patients with 24,908 notes across 69 note types. Six additional patients were excluded from model development and validation because their available notes contained no cancer-relevant clinical content (e.g., administrative telephone encounters only), yielding a final development cohort of 579 patients with 24,887 notes across 69 note types for overall metastasis analysis. Three additional patients with unconfirmed brain metastasis registry records were excluded from the brain/CNS metastasis analysis, yielding a cohort of 576 patients. The full cohort selection process is illustrated in Figure 1. Demographic characteristics, following initial data extraction and cohort selection, are described in Table 1.

**Fig. 1:**
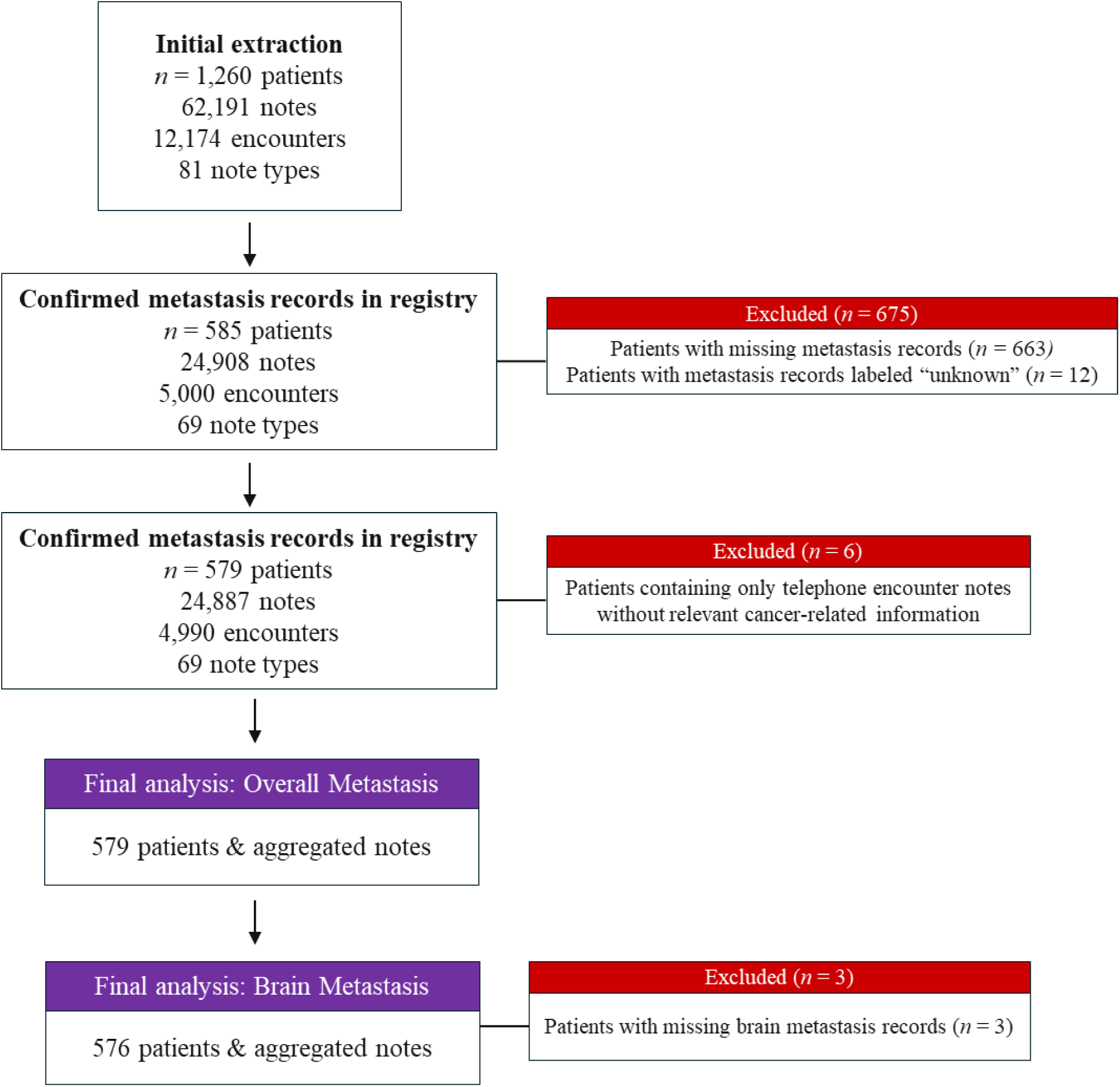
Flowchart of selection of Cohort 1 following initial data extraction.

**Fig. 2:**
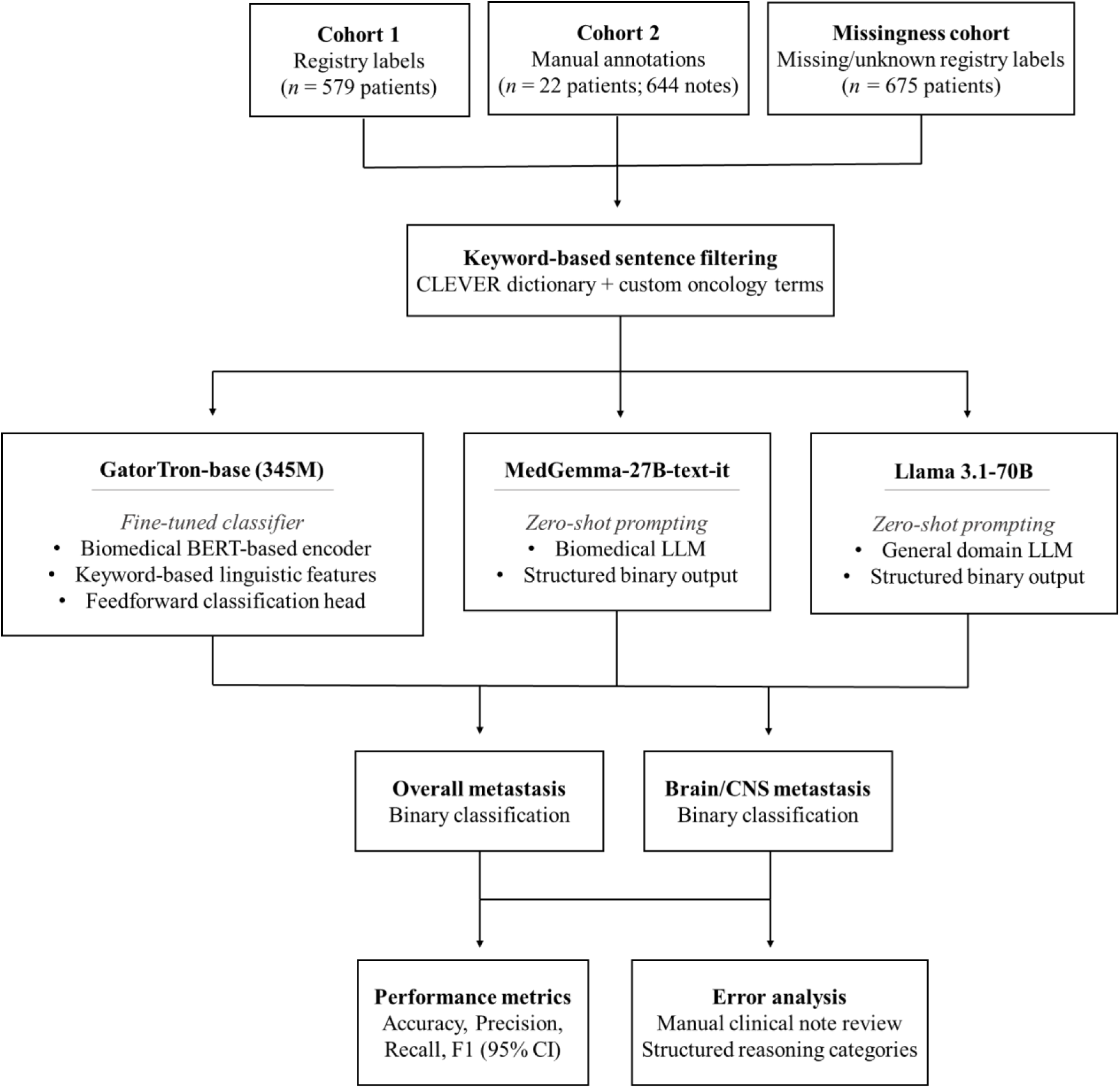
Note preprocessing and model inference pipeline.

**Table 1:**
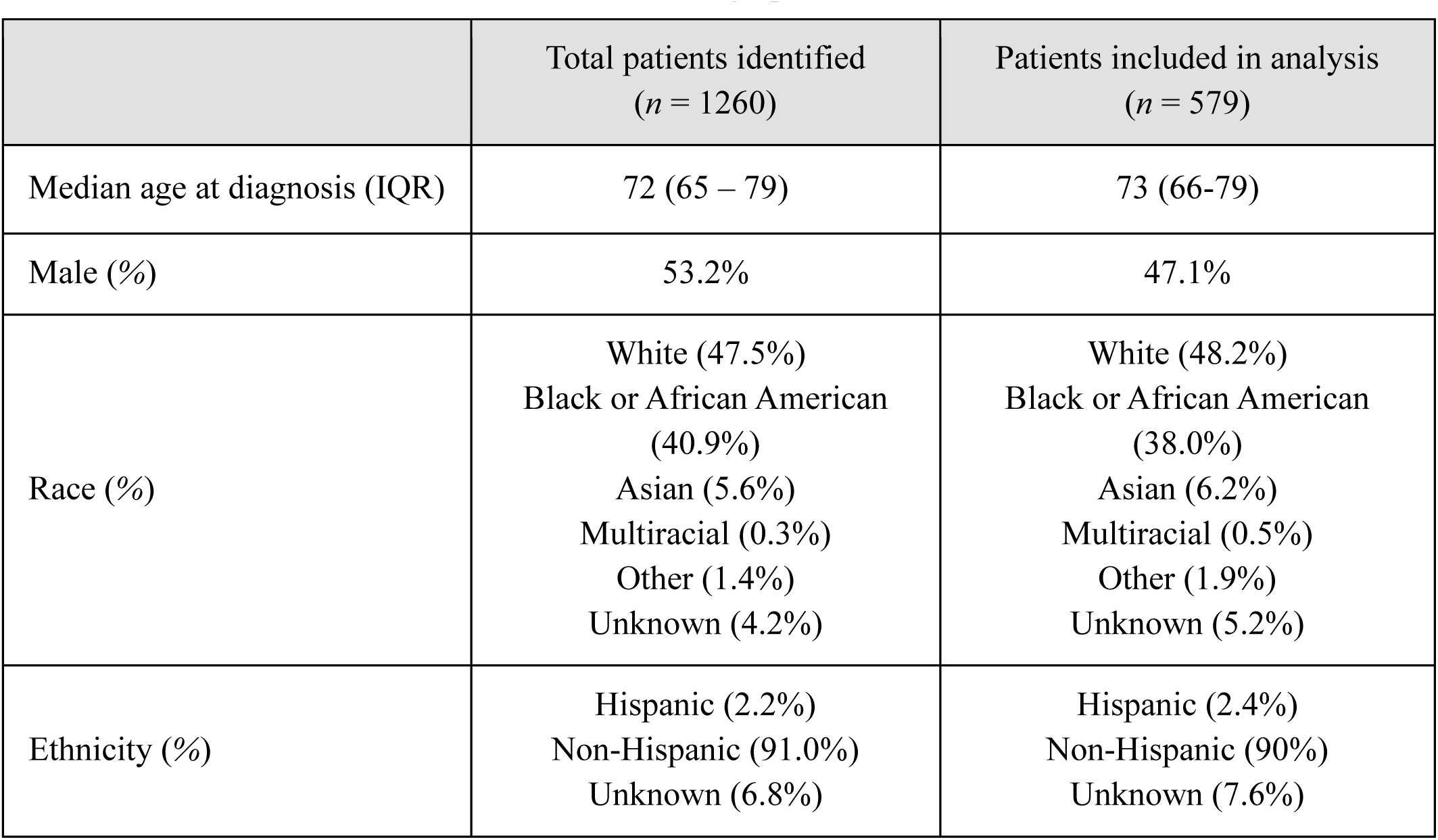
Cohort 1 Demographic Characteristics.

**Table 2:** Overall Cohort Characteristics.

|  | Cohort 1:<br>Primary Registry-Linked | Cohort 2:<br>Temporal Validation |
| --- | --- | --- |
| Reference Standard | Cancer registry | Manual annotation |
| Documentation period | 2023–2025 | 2010–2021 |
| Total NSCLC patients identified, $n$ | 1,260 | 22 |
| Patients with missing/unknown registry metastasis records, $n$ (%) | 675 (54%) | No missingness |
| Patients included in analysis, $n$ | 579 (overall)<br>576 (brain/CNS) | 22 |
| Clinical notes included in analysis, $n$ | 24,887 | 644 |
| Note types | Heterogeneous (69 types) | Radiology only |
| Patients with overall metastasis, $n$ (%) | 239 (41%) | 22 (100%) |
| Patients with brain/CNS metastasis, $n$ (%) | 89 (15%) | 8 (36%) |

#### Reference standard

We used the Winship Cancer Registry as our reference standard, which reports site-specific metastasis (liver, brain, bone, lung, distant lymph nodes, other distant site). Using these site-specific fields, we created our binary outcome label to identify overall metastasis. This label, comprehensive of all metastasis sites, was derived as follows: patients with at least one site-specific positive metastasis record were classified as metastasis-positive, patients with all-negative records were classified as metastasis-negative, and patients with at least one missing (*n* = 663) or unknown (*n* = 12) field, in the absence of a confirmed positive, were classified as metastasis unknown and excluded from model development. The registry brain metastasis field was used directly as the reference standard analysis for brain metastasis analysis.

#### Note inclusion window

Because registry metastasis fields primarily capture disease status at initial diagnosis, clinical note inclusion was restricted to a window spanning 30 days before through 30 days after each patient’s formal cancer diagnosis date. This window maximized temporal correspondence between note content and registry labels. All note types within this window were eligible, including radiology reports, discharge summaries, progress notes, consultation notes, provider communication, nursing documentation, and other note types. The full distribution of notes across all note types is detailed in Supplemental Table S1.

#### Cohort 2: Independent Temporal Validation Cohort

To assess model generalizability across an earlier documentation era, we assembled an independent cohort of 22 NSCLC patients whose records were extracted retrospectively from two completed randomized clinical trials (RCTs) conducted between 2010 and 2021. Trial enrollment required systematic imaging, resulting in dense radiology documentation in the historical records of these patients. A total of 644 radiology notes, including CT, MRI, X-ray, and other imaging reports were extracted. Notes were not restricted to a diagnosis-adjacent window, and registry records were not used as the reference standard, since they did not capture metastatic progression documented longitudinally. Manual note-level annotations served as the reference standard labels for this cohort (*Section D*). All models were applied to this cohort without retraining to serve as further external validation.

#### Missingness-Recovery Dataset

The 675 patients excluded from primary model development, due to at least one missing or unknown registry metastasis record, represented 54% of the initial Cohort 1 extraction pool. 12 of these patients had unknown brain metastasis records. Notes from these patients were retained for a downstream exploratory analysis. This dataset was used solely to assess whether our methods could recover structured metastasis information absent from registry fields. No reference standard was available for this group, and model predictions were qualitatively reviewed by selecting a small random sample of patients and manually assessing their clinical notes, which were aggregated to the patient level. (*Section H*). This analysis is reported as a feasibility exercise and not as a primary performance evaluation.

### C. Clinical Note Preprocessing

All clinical notes were preprocessed through a keyword-based filtering pipeline prior to model input, designed to reduce noise while retaining clinically relevant content. Keywords were drawn from the publicly available CLEVER dictionary, which contains clinical synonyms for oncology-relevant terms, supplemented by custom oncology-specific terms developed by the study team [30]. Representative CLEVER terms included “metastasis,” “widespread disease,” and “stage IV”, among other words and phrases. Custom additions included “high grade” and “malignant mass”, in addition to other words. All keywords used for preprocessing are available in Supplemental Table S2. For notes containing at least one keyword match, only sentences containing keywords were retained. For notes without any match, the full note text was preserved. This step was applied strictly as a noise-reduction measure to eliminate spans irrelevant to the patient’s disease course. In Cohort 1, all notes from each patient were aggregated at the patient level before preprocessing and model input, consistent with the patient-level registry reference standard. In Cohort 2, notes were processed and inputted individually.

### D. Manual Annotation of Cohort 2

Two trained abstractors, both of whom have a background in public health and biomedical informatics, independently annotated radiology notes from Cohort 2 under the supervision of a board-certified oncologist. A standardized schema targeted two binary note-level labels: (1) overall metastasis present versus absent, and (2) CNS metastasis present versus absent. Annotators were instructed to label both explicit evidence (e.g., “brain metastases confirmed on MRI”) and contextually implied evidence strongly suggestive of current metastatic disease, such as imaging findings or treatment references indicating metastatic involvement. Disagreements were resolved by discussion or oncologist adjudication. Annotation proceeded in four iterative rounds of increasing size (*n* = 10, 100, 200, and 334 notes), with inter-annotator agreement assessed and discrepancies reconciled between rounds. Agreement was quantified using pairwise Cohen’s κ. Final κ values, across all rounds of annotation, were 0.93 for overall metastasis and 0.88 for CNS metastasis, indicating high agreement (Table 3). Final labels were assigned based on unanimous consensus or oncologist suggestion.

**Table 3:** Inter-annotator agreement for Cohort 2.

| Annotation Round | Cohen's $\kappa$ | |
| --- | --- | --- |
|  | Overall Metastasis | CNS Metastasis |
| Round 1 ($n = 10$) | 1 | 1 |
| Round 2 ($n = 100$) | 0.92 | 0.80 |
| Round 3 ($n = 200$) | 0.92 | 0.85 |
| Round 4 ($n = 334$) | 0.93 | 0.91 |

### E. Model Development and Inference

We evaluated three pretrained large language models representing distinct architectures: (1) GatorTron-base, (2) MedGemma-27B-text-it, and (3) Llama 3.1-70B [31–33].

*GatorTron* is a transformer encoder pretrained on more than 90 billion words of deidentified clinical text, biomedical literature, and other English text [31]. We began by benchmarking the non-fine-tuned GatorTron-base model (345M parameters) for binary classification of overall metastasis on a subset of Cohort 1. This initial benchmarking yielded random chance predictions (F1=0.46), indicating a need for further fine-tuning on our institutional cohort [34]. We fine-tuned two separate GatorTron-base models for binary classification of overall metastasis (80% train, *n* = 463; 20% test, *n* = 116) and brain/CNS metastasis (80% train, *n* = 461; 20% test, *n* = 115). To mitigate label noise arising ambiguous phrasing in clinical note text, training incorporated extraction of key linguistic features in the clinical text. Firstly, a list of metastasis-relevant terms, from the CLEVER dictionary, was used to capture mentions of metastasis presence. For fine-tuning of brain/CNS metastasis classification, these terms were expanded to include brain/CNS site-specific verbiage. Next, NegEx-based negation detection was implemented handle negated metastasis mentions [35]. Finally, a curated list of uncertainty expressions (e.g., “suspicious for metastasis,” “may represent metastasis”) was implemented handle unconfirmed metastasis mentions. These features were encoded as binary indicators. The full list of metastasis-relevant terms used during fine-tuning of each classification task are included in Supplemental Table S3.

Notes in the 80% train set were further partitioned into an 80% train set (overall metastasis: *n* = 370, brain/CNS metastasis: *n* = 369) and 20% validation set (overall metastasis: *n* = 93, brain/CNS metastasis: *n* = 92), stratified based on the binary outcome label. Clinical notes were tokenized and split into overlapping 512-token chunks with a stride of 128 tokens to accommodate long notes. We trained a custom classifier head that concatenated the three linguistic features, described above, into a single feature vector. This feature vector, along with the text representation generated by GatorTron, were inputted into a feedforward neural network, which incorporated contextual and rule-based information to generate a binary prediction of metastasis presence. The 20% validation set was used to identify the optimal probability threshold for metastasis classification by evaluating thresholds between 0 to 1 and selecting the probability value that optimized the F1 score per patient. Final inference was conducted on the 20% hold-out test set (overall metastasis: *n* = 116, brain/CNS metastasis: *n* = 115). Final patient-level predictions were assigned based on the maximum probability predicted across chunks corresponding to each patient’s note. Predictions on Cohort 2 were generated using the final fine-tuned model without further retraining.

*MedGemma-27B-text-it* is a biomedical variant of Google’s Gemma model, optimized for medical text and image understanding [32]. We evaluated the 27 billion parameter instruction-tuned text model in a zero-shot setting using structured prompts that instructed the model to classify whether the provided note text documented the target condition and to return a constrained binary response (“Metastasis: YES” or “Metastasis: NO”). Greedy decoding with a fixed maximum output length was used to minimize output variability. Prompt templates are provided in Supplemental Materials 1 and 2.

*Llama 3.1-70B-Instruct* is a general-domain open-source LLM developed by Meta, trained on a large-scale corpus of general text and optimized for instruction-following [36]. Unlike MedGemma, it was not specifically pretrained on clinical or biomedical text. Llama3 was evaluated using an identical zero-shot prompt structure to MedGemma, decoding configuration, and constrained binary output format as MedGemma for both extraction tasks.

Given these differences in level of training and fine-tuning involved in LLM evaluation, this study design enabled assessment of distinct NLP strategies for metastasis classification rather than a direct comparison based only on underlying LLM architectures. All model weights were obtained from Hugging Face and deployed locally on an institutional, HIPAA-secure computing infrastructure [37–39]. All analyses were implemented in Python 3.10. All code is available on GitHub [40].

### F. Data Partitioning and Model Evaluation

GatorTron fine-tuning and testing were conducted on Cohort 1. MedGemma and Llama3 did not undergo fine-tuning and were instead evaluated under a zero-shot prompt setting. Cohort 1 evaluations for all models were conducted on the 20% test set used in the evaluation of the fine-tuned GatorTron model. Therefore, final Cohort 1 performance, for all models, were calculated based on model predictions generated for the same 20% test set of the total cohort (overall metastasis: *n* = 116, brain metastasis: *n* = 115).

Cohort 2 served as an independent out-of-sample test set for all three models. Since note-level manual annotations served as the reference standard for Cohort 2, model evaluation was conducted at both the note-level (*n* = 644) and the patient-level (*n* = 22). Patient-level reference standards were derived by aggregating note-level annotations, such that if any note of a patient is labeled metastasis positive, the patient-level label is metastasis positive. For each model, cohort, and prediction level, we computed accuracy, precision, recall, and F1 score. Ninety-five percent confidence intervals were estimated using bootstrapped resampling with 10,000 iterations. All metrics were reported separately for the overall metastasis and brain/CNS metastasis tasks.

### G. Error Analysis

For the best and poorest performing models on each extraction task in the development cohort, all misclassified cases were examined qualitatively. Clinical notes from each error were manually reviewed and assigned to one of four pre-specified error categories: (1) incorrect model prediction despite obvious note content, (2) ambiguous clinical text, in which metastatic status was uncertain or unconfirmed by imaging, laboratory testing, or biopsy, (3) correct model prediction with an erroneous registry label, or (4) missing clinical text, in which relevant documentation was likely absent from the available note set. Category definitions and representative examples are provided in Table 4.

**Table 4:** Structured Reasoning Categories used in Qualitative Analysis of Prediction Errors.

| Error Category | Description | Example |
| --- | --- | --- |
| Incorrect Prediction | The model assigned an incorrect prediction despite clear indication of positive or negative metastasis presence in the clinical note text. | “Suspected metastasis confirmed to be benign meningioma.”<br>In this case, the note explicitly confirms negative metastasis, but the model predicts a positive case. |
| Ambiguous text | The clinical note text has unclear confirmation of metastasis presence. The note expresses uncertainty of metastasis presence without documented confirmation via imaging, lab test, or biopsy.<br>The clinical note text may also indicate suspicion of an additional primary cancer, which the model predicts as metastasis. | “Brain metastasis cannot be ruled out.”<br>“There are several malignant nodular lesions.”<br>If no imaging, lab test, or biopsy confirmation is documented, the model may predict the incorrect label. |
| Correct prediction and incorrect registry label | In some cases of false positive predictions, the available clinical note demonstrates clear mentions of metastasis presence, either indicating an error in the registry label or an update in the patient’s cancer diagnosis not reflected in the registry. | “Biopsy results confirm osseous mets.”<br>If this confirmation is documented around the time of initial cancer diagnosis but is not reflected in the cancer registry, this may reflect an error in the registry label. |
| Missing clinical text | In some cases of false negative predictions, the available clinical note does not clearly state any mention of metastasis. Therefore, rather than a failure of the model, these errors indicate the possible presence of additional notes, unavailable to us, which may have included metastasis information. | If the registry indicates metastasis to a specific site, but there is no language in the clinical note relevant to metastasis, metastasis testing, or even mention of the site, the model prediction will not align with the registry label. |

### I. Exploratory Downstream Missingness-Recovery Analysis

The best-performing model for each extraction task was applied to the 675 patients in the missingness-recovery dataset. This analysis was conducted to evaluate the feasibility of leveraging our models to recover metastasis information from narrative text. To assess prediction validity, random samples of 10 predicted-positive and 10 predicted-negative cases were drawn, and their notes were manually reviewed and labeled prior to viewing model predictions to ensure unbiased assessment. Concordance between manual labels and model predictions was then evaluated. Given the small validation sample, findings are not definitive, but rather exploratory.

## Results

### A. Cohort Characteristics

Among 1,260 patients with NSCLC identified from the Winship Cancer Registry and linked to clinical notes, restricted to analytic cases diagnosed and/or treated at Emory, metastasis status was missing or recorded as unknown in 54% of patients. Cohort 1 included 579 patients with complete registry metastasis data and 24,898 clinical notes; 41% had metastasis at any site and 15% had brain metastasis at diagnosis. Cohort 2 included 22 patients and 644 radiology notes. All patients had metastasis at any site, and 36% had CNS metastasis. At the note level, 51% of notes were positive for metastasis and 11% for CNS metastasis.

### B. Model Performance: Overall Metastasis Extraction

All models demonstrated strong performance (Table 5). In Cohort 1, MedGemma demonstrated the best performance (F1=0.80), followed by Llama3 (F1=0.76), and fine-tuned GatorTron (F1=0.72) In Cohort 2, all models achieved high patient-level performance, which was expected given the availability of dense radiological documentation and the presence of metastasis in all patients. At the note level, MedGemma performed best (F1=0.93), followed by Llama3 (F1=0.88) and GatorTron (F1=0.76). The optimal probability threshold determined for overall metastasis classification was 0.58.

**Table 5:**
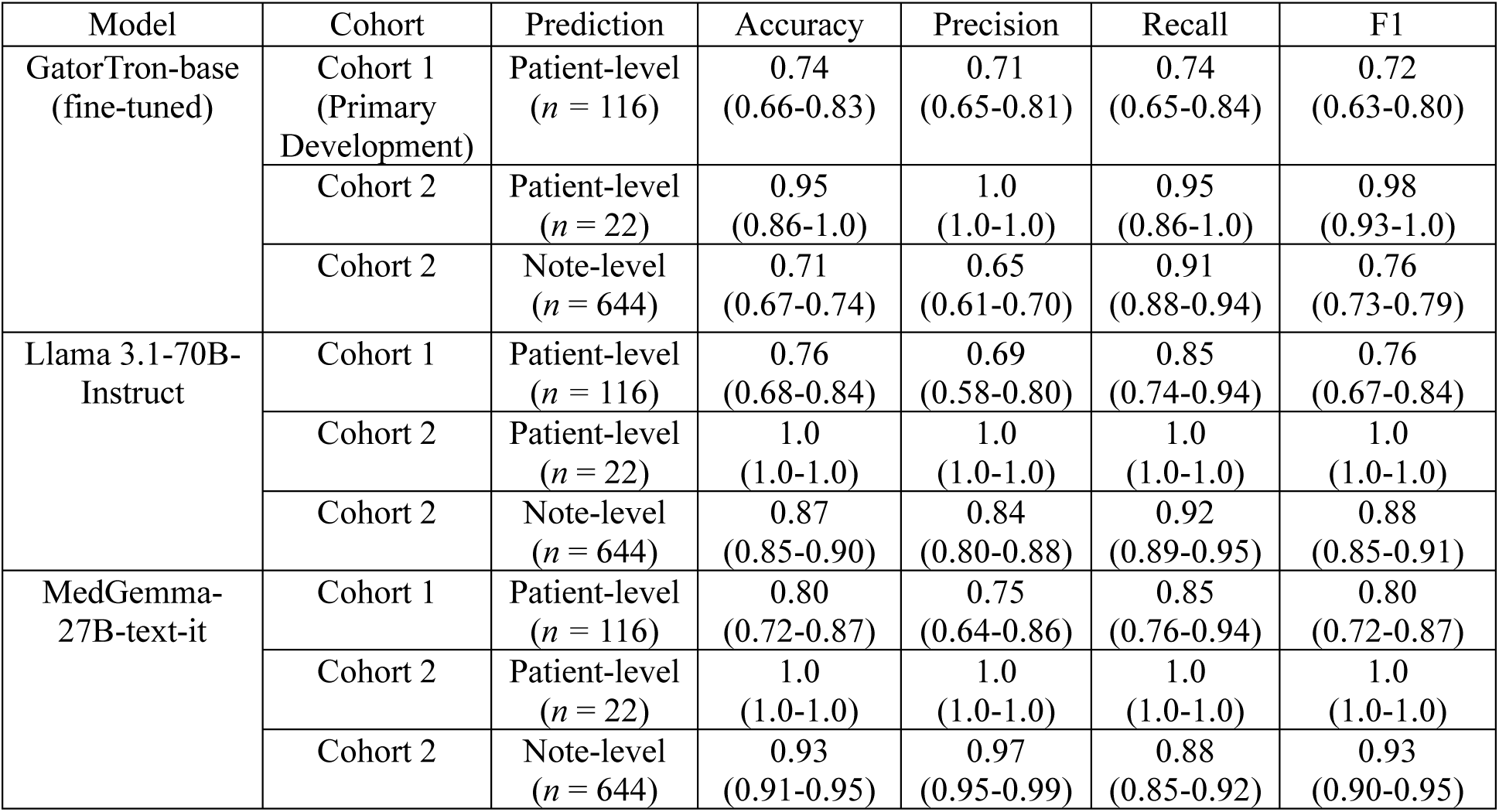
Performance Metrics and 95% Confidence Intervals for Overall Metastasis Prediction.

**Table 6:** Performance Metrics and 95% Confidence Intervals for Brain/CNS Metastasis Prediction.

| Model | Cohort | Prediction | Accuracy | Precision | Recall | F1 |
| --- | --- | --- | --- | --- | --- | --- |
| GatorTron-base<br>(fine-tuned) | Cohort 1<br>(Primary Development) | Patient-level<br>($n = 115$) | 0.88<br>(0.80-0.96) | 0.50<br>(0.38-0.64) | 0.50<br>(0.26-0.71) | 0.50<br>(0.34-0.66) |
| | Cohort 2 | Patient-level<br>($n = 22$) | 0.73<br>(0.55-0.91) | 0.57<br>(0.30-0.82) | 1.0<br>(1.0-1.0) | 0.72<br>(0.46-0.90) |
| | Cohort 2 | Note-level<br>($n = 644$) | 0.81<br>(0.78-0.84) | 0.35<br>(0.28-0.42) | 0.83<br>(0.75-0.92) | 0.50<br>(0.42-0.67) |
| Llama 3.1-70B-Instruct | Cohort 1 | Patient-level<br>($n = 115$) | 0.96<br>(0.91-0.99) | 0.91<br>(0.69-1.0) | 0.71<br>(0.46-0.94) | 0.79<br>(0.59-0.96) |
| | Cohort 2 | Patient-level<br>($n = 22$) | 0.95<br>(0.86-1.0) | 0.88<br>(0.62-1.0) | 1.0<br>(1.0-1.0) | 0.93<br>(0.77-1.0) |
| | Cohort 2 | Note-level<br>($n = 644$) | 0.96<br>(0.95-0.98) | 0.82<br>(0.73-0.90) | 0.90<br>(0.82-0.97) | 0.86<br>(0.79-0.91) |
| MedGemma-27B-text-it | Cohort 1 | Patient-level<br>($n = 115$) | 0.95<br>(0.90-0.98) | 0.83<br>(0.58-1.0) | 0.72<br>(0.45-0.94) | 0.76<br>(0.54-0.92) |
| | Cohort 2 | Patient-level<br>($n = 22$) | 0.82<br>(0.74-0.95) | 0.83<br>(0.43-1.0) | 0.63<br>(0.25-1.0) | 0.70<br>(0.33-0.93) |
| | Cohort 2 | Note-level<br>($n = 644$) | 0.97<br>(0.95-0.98) | 0.83<br>(0.75-0.92) | 0.89<br>(0.81-0.96) | 0.86<br>(0.79-0.91) |

### C. Model Performance: Brain/CNS Metastasis Extraction

In Cohort 1, Llama3 achieved the highest performance (F1=0.79), followed by MedGemma (F1=0.76). GatorTron achieved comparably lower performance in this cohort (F1=0.50). In Cohort 2, Llama3 again performed best (patient-level F1=0.93; note-level F1=0.86). MedGemma achieved comparable note-level performance (F1=0.86) with slightly lower patient-level performance (F1=0.70). GatorTron achieved high patient-level performance (F1=0.72) with relatively low note-level performance (F1=0.50). The optimal probability threshold determined for overall metastasis classification was 0.40.

### D. Error Analysis

Prediction errors from the best and poorest performing models for each classification task in the held-out test set of Cohort 1 (overall metastasis [*n* = 116], brain/CNS metastasis [*n* = 115]) were qualitatively analyzed and assigned to a category of reasoning. Corresponding predictions from the other two models were assessed in parallel.

### Overall Metastasis Prediction

#### GatorTron

GatorTron generated 14 false negative predictions and 16 false positive predictions. Less than half of the errors generated (*n* = 13) were due to incorrect model errors, all of which were predicted correctly by MedGemma and Llama3. The primary reason for other errors was ambiguous phrasing in the clinical text (*n* = 10), in which clinical documentation indicated “suspicious,” “concerning,” or “probable” metastasis without clear confirmation through lab test results. Another reason was due to the model correctly generating positive predictions (*n* = 4), despite a metastasis negative registry label. In these cases, patient notes explicitly mentioned diagnosis of metastatic cancer, which were not reflected in the registry but identified by our model. Finally, multiple false negative cases (*n* = 3) were due to missing clinical text. A detailed error analysis is provided in Supplemental Table S4.

#### MedGemma

For prediction of overall metastasis, MedGemma generated 8 false negative predictions and 15 false positive predictions. In 9 instances, the model generated correct, positive predictions despite conflicting registry records. Errors also occurred due to missing clinical text (*n* = 6) and ambiguous clinical text (*n* = 6) (four false positive and two false negative predictions). Finally, the model produced one false positive prediction and one false negative prediction, both categorized as incorrect and predicted correctly by Llama3 and GatorTron. A detailed error analysis is provided in Supplemental Table S5.

### Brain/CNS Metastasis Prediction

#### GatorTron

For brain/CNS metastasis, GatorTron generated 7 false negative predictions and 7 false negative predictions. Most errors were caused by incorrect predictions (*n* = 10), all of which were all predicted correctly by MedGemma and Llama3. Other errors were due to missing text (*n* = 3) and ambiguous text (*n* = 1). A detailed error analysis is provided in Supplemental Table S6.

#### Llama3

Llama3 generated 4 false negative predictions due to missing clinical text and one false positive prediction, which was determined to be incorrect. The incorrect false positive prediction was predicted correctly by MedGemma and GatorTron. A detailed error analysis is provided in Supplemental Table S7.

### E. Exploratory Missingness-Recovery Analysis

We evaluated whether models could recover metastasis information among 675 patients with at least one missing or unknown registry label (including 12 with unknown brain metastasis status). Using the best-performing model for overall metastasis, MedGemma classified 302/675 patients (44.7%) as metastasis-positive, closely resembling the prevalence of metastasis in Cohort 1 (41%). To assess validity, a random sample of 20 cases (10 predicted positive and 10 predicted negative) were manually annotated prior to viewing model results. Agreement with model predictions yielded accuracy = 0.90 and F1=0.89. Two false positive errors were identified in prediction of overall metastasis due to several ambiguous phrases regarding suspicion of metastasis without clear confirmation. For brain/CNS metastasis, all 12 patients with unknown registry status were manually annotated as negative and correctly classified by Llama3.

## Discussion

LLM-based methods demonstrated strong performance for extracting overall and CNS metastasis from real-world NSCLC clinical notes, with performance varying by task and evaluation setting. Additional exploratory analyses suggest that clinically meaningful information about metastasis can be recovered from narrative documentation when structured registry data are missing. These findings highlight a core limitation of current oncology data systems. Cancer registries, while essential, are temporally limited and incomplete, whereas clinical notes can provide longitudinal, detailed documentation of disease evolution. LLMs offer a practical mechanism to extract clinically relevant information embedded in routine documentation and complement structured data.

Earlier studies have largely focused on single cohorts or limited note types, particularly radiology reports and discharge summaries.^24,25^ By leveraging a registry-linked cohort with heterogeneous oncology documentation, we provide a more realistic evaluation of real-world performance. In addition, few studies have directly compared clinically pretrained encoder models with large instruction-tuned generative models on the same task.^8,11,12^ In our study, baseline performance metrics demonstrated that the generative models (Llama3 and MedGemma) achieved higher performance in classifying both overall and brain/CNS metastasis presence, respectively, compared to the fine-tuned GatorTron model. Upon further inspection of model errors, generative models and fine-tuned encoder models appeared to serve complementary strengths, where certain prediction errors made by one model were correctly predicted by another. Therefore, these findings do not necessarily indicate the overall superiority of one model architecture over another for metastasis classification tasks, given the limitations present in cancer registry records. Fine-tuned encoder models show promise for classifying metastasis presence and identifying potentially inaccurate registry records based on clinical note text; however, they may be sensitive to discrepancies present in their input data. Instruction-tuned generative models, even in a zero-shot prompt setting, demonstrate holistic understanding of clinical note text without this limitation, which may explain the differences in model performance in this study.

In this multi-cohort evaluation, we make three principal observations. First, metastasis information was substantially incomplete even within a cancer registry-linked dataset, with more than half of patients lacking definitive structured metastasis labels. Second, model performance varied, with the instruction-tuned generative models demonstrating holistic understanding of complex clinical language and the fine-tuned encoder model demonstrating strong classification performance but likely hindered by potentially inaccurate registry labels during training. Third, LLMs appear adept at correcting inaccurate registry records and recovering missing data.

Our first observation in this study is the high prevalence of missing data in structured metastasis labels, representing 54% of the patients identified in our initial extraction of Cohort 1. This study extends prior clinical NLP work in important ways. While cancer registries serve as the gold standard for cancer surveillance, future studies may seek to use more curated real-world datasets for more robust model fine-tuning. Though inaccuracies and missingness in the cancer registry may have inhibited the fine-tuned encoder model from more accurately distinguishing between positive and negative metastasis cases, our model still demonstrated the ability to correct and inaccurate data based on clinical text documentation.

Our second observation in this study is that model performance patterns underscore that no single approach is uniformly optimal. Encoder models align well with structured labels; however, discrepancies in registry records may influence model outputs. In contrast, generative models appear more robust to heterogeneous, implicit, and context-dependent clinical language, which is particularly relevant for CNS metastasis, where documentation is often indirect and radiology-driven. This difference in performance may be attributed to the ability of generative models to better identify nuanced and complex site-specific language used in the documentation of brain/CNS without influence of structured registry labels. Furthermore, GatorTron’s prediction of brain/CNS metastasis may have been inhibited by the class imbalance in brain/CNS metastasis cases during model fine-tuning, whereas this class imbalance was not impactful in a zero-shot prompting setting. These results support a use-case-specific deployment strategy rather than a single-model solution.

Our third observation in this study is that LLM-based extraction appears to be adept at correcting and recovering incomplete metastasis data in structured registry fields. Our error analysis indicated that many discrepancies were due to data limitations in reference labels or available clinical documentation rather than explicit model failure. Differences in registry labels compared to clinical documentation, missing clinical text, and ambiguity in clinical language accounted for most errors. In several cases, model predictions were clinically plausible despite conflicting with registry labels, underscoring the imperfect nature of structured ground truth in oncology datasets. Both the error analysis and exploratory missingness recovery analysis provide an initial signal that NLP may augment registry accuracy and completeness. The error analysis identified several positive metastasis cases that were not reflected in the registry labels, despite clinical documentation near the time of initial diagnosis. The exploratory missingness recovery analysis demonstrated high accuracy recovering missing metastasis labels, based on manual review of a subset of notes. While preliminary, this suggests that clinically relevant signals embedded in narrative documentation can be systematically recovered. These findings have direct clinical and operational implications. NLP-based extraction can support cancer registry workflows by prioritizing high-yield cases and improving completeness. It may also enable more accurate cohort construction and clinical trial screening, where metastasis status, particularly CNS involvement, is a key eligibility criterion.

Our study has several limitations. First, Cohort 1 (the primary development cohort) relied on cancer registry labels that contain limitations, including accuracy of label and inability to capture metastatic progression. Thus, discordance between registry labels and model predictions does not necessarily indicate model error, since the reference standard itself may be imperfect. Furthermore, there is a possibility that, though documentation spanned one month before until one month after initial diagnosis, metastasis cases diagnosed within one month after initial cancer diagnosis were not updated in the registry. Second, Cohort 2 was small and derived from clinical trial populations with dense radiologic follow-up, which may limit generalizability to routine care settings. Additionally, the documentation style in this cohort of notes may have demonstrated meaningful variation from the Cohort 1 notes used for primary model development. Thus, this discrepancy may have hindered the performance of the fine-tuned model in Cohort 2. Third, the use of different reference standards across cohorts (registry-based at the patient level in Cohort 1 vs manual note-level annotation in Cohort 2) complicates direct comparison, particularly given differences in temporal scope and definition of CNS metastasis. Fourth, models of differing underlying architectures were evaluated in distinct ways, wherein GatorTron was fine-tuned, while generative models were evaluated in a zero-shot setting. Fifth, the keyword-based preprocessing strategy may have excluded relevant contextual information, potentially limiting sensitivity to implicitly expressed metastasis. Sixth, the study was conducted at a single institution, and broader external validation is needed. Finally, the missing recovery analysis was exploratory and based on a small sample; a larger scale validation should be conducted to further evaluate the potential of recovering missing metastasis records.

## Conclusion

In this study the instruction-tuned generative models, evaluated in a zero-shot setting, performed best at classifying metastasis. The high degree of missing metastasis data, even within a cancer registry-linked cohort, underscores a key limitation of structured oncology data and highlights the value of note-based ascertainment. Exploratory analyses further suggest that metastasis information documented in clinical narratives can be partially recovered when structured fields are incomplete. These findings support the use of NLP to complement cancer registry data for metastasis ascertainment, cohort identification, and trial screening, while requiring further validation before broader use.

## Data Availability

The datasets analyzed in this study are not publicly available due to institutional policies regarding HIPAA (Health Insurance Portability and Accountability Act) protected medical files.

## Acknowledgements

Patient cohort identification was conducted by the Winship Cancer Institute’s Data and Technology Applications Shared Resource (WinDATA). Clinical note extraction for select patients was conducted by Emory University’s Medical Informatics and AI Core team.

## Author Contributions

Conceptualization: SBo, RBP

Data curation: SBa, RC, SBo

Formal Analysis: SBa, KC

Funding acquisition: SBo

Investigation: SBa, SBo

Methodology: SBa, SBo, KC

Project administration: SBa, SBo

Supervision: SBo, RBP, JDW, AS

Validation: SBo, KC

Visualization: SBa

Wiring-original draft: SBa, SBo

Writing-review & editing: SBa, SBo, KC, RC, RBP, JDW, MR, DS

## Abbreviations

CNS: central nervous system
EHR: electronic health record
LLM: large language model
NLP: natural language processing
NSCLC: non-small cell lung cancer
RCT: randomized controlled trial
ML: machine learning

