## Supplemental Materials for "Metastasis Extraction from NSCLC Clinical Notes: A Retrospective Comparative Evaluation of Large Language Model-Based Classification"

Table S1. Distribution of Note Types in Each Cohort

Table S2. Keywords Used in Note Preprocessing Pipeline

Table S3. Keywords Used for Feature Extraction During GatorTron Fine-tuning

Supplemental Material 1. Prompt Template used for Prediction of Overall Metastasis

Supplemental Material 2. Prompt Template used for Prediction of Brain/CNS Metastasis

Table S4. GatorTron Error Analysis for Overall Metastasis Prediction

Table S5. MedGemma Error Analysis for Overall Metastasis Prediction

Table S6. GatorTron Error Analysis for Brain/CNS Metastasis Prediction

Table S7. Llama3 Error Analysis for Brain/CNS Metastasis Prediction

### Table S1. Distribution of Note Types in Each Cohort

| Cohort | Note Type | # of notes, % of all notes |
|---|---|---|
| Cohort 1 | Progress Notes | 7341, 29.5% |
| | Care Plan | 3795, 15.2% |
| | Telephone Encounter | 3678, 14.8% |
| | Nursing Note | 2426, 9.7% |
| | Consults | 1027, 4.1% |
| | History & Progress | 740, 3.0% |
| | ED Notes | 636, 2.6% |
| | Addendum Note | 432, 1.7% |
| | Anesthesia Procedure Notes | 428, 1.7% |
| | PatientPass | 426, 1.7% |
| | Anesthesia Preprocedure Evaluation | 385, 1.5% |
| | Procedures | 376, 1.5% |
| | Anesthesia Postprocedure Evaluation | 346, 1.4% |
| | Discharge Summary | 304, 1.2% |
| | ED Provider Notes | 283, 1.1% |
| | Discharge Instructions | 242, 1.0% |
| | Perioperative Nursing Note | 234, 0.9% |
| | Significant Event | 223, 0.9% |
| | ED Triage Notes | 206, 0.8% |
| | Provider Documentation Clarification | 137, 0.6% |
| | Preprocedure Instructions | 130, 0.5% |
| | Radiation Oncology CT Sim/CTP | 106, 0.4% |
| | Result Encounter Note | 98, 0.4% |
| | Radiation Oncology Consult | 94, 0.4% |
| | Op Note | 93, 0.4% |
| | Discharge Plan Summary | 87, 0.3% |
| | Patient Instructions | 78, 0.3% |
| | Post-Procedure Note | 66, 0.3% |
| | Brief Op Note | 56, 0.2% |
| | Student | 49, 0.2% |
| | Sedation Documentation | 48, 0.2% |
| | ED Procedure Note | 33, 0.1% |
| | Assessment & Plan Note | 32, 0.1% |
| | Malnutrition | 49, 0.2% |
| | Hospital Course | 23, 0.1% |

| | | |
|---|---|---|
| | Patient Communication | 23, 0.1% |
| | Discharge Instructions | 39, 0.2% |
| | Case and Service Plan | 21, 0.1% |
| | Unmapped External Note | 16, 0.1% |
| | Pre-Procedure Note, Pre-Sedation Documentation, ED Addendum Note, Family Meeting Note, Radiation Completion Note, Transfer of Care, Payer Review Note, Preadmission Screening, Infection Prevention, Code Documentation, Multidisciplinary Conference Note, Advance Care Planning, Individualized Overall Plan, OR Surgeon, Radiation Planning Notes, Committee Review, Case Communication, Dental Procedure Details, OR PostOp, Subjective & Objective, Downtime Event Note | 81, <0.1% |
| Cohort 2 | CT Scan | 383, 59.4% |
| | MRI Scan | 115, 17.8% |
| | X-Ray Scan | 110, 17.1% |
| | PET/CT Scan | 17, 2.6% |
| | IR Scan | 8, 1.2% |
| | NM Scan | 6, 0.9% |
| | CTA Scan | 3, 0.5% |
| | MRA Scan | 2, 0.3% |

### Table S2. Keywords Used in Note Preprocessing Pipeline

| | |
|---|---|
| CLEVER Dictionary | metastasis, metasasis, metastisis, metatasis metastases, metasases, metatases, metastatic, metastitic, metastic, metatatic, metastasize, metastatize, metastisize, metastasize, metastasized, metastised, metastatized,, metastasized, metastasizing, metastisizing, metatasizing<br><br>mets, mtas, msts, met's, met dz, met dis, met disease, met ca, m. ca, mca, mCA, MCA |

| | |
|---|---|
| | mCRC, m-CRC, mCRCa, mBC, m-BC, mBrCa, adv dz, adv dis, adv disease, adv ca, adv cancer<br><br>bone metastasis, bony metastasis, bone mets, bony mets, osseous metastasis, oss mets, osseus metastases lung metastasis, pulmonary metastasis, lung mets, pulm mets, pulmonary mets, liver metastasis, hepatic metastasis, liver mets, hepatic mets, hep mets<br>brain metastasis, cerebral metastasis, brain mets, CNS mets, cereb mets, intracranial mets<br>leptomeningeal metastasis, meningeal metastasis, leptomeningeal mets, meningeal mets, lymph node metastasis, nodal metastasis, LN mets, lymphatic mets, LN involvement, adrenal metastasis, adrenal mets<br>peritoneal metastasis, peritoneal mets, carcinomatosis, peritoneal carcinomatosis<br>pleural metastasis, pleural mets, pleural involvement, pleural spread<br>cutaneous metastasis, skin metastasis, cutaneous mets, skin mets<br><br>distant metastasis, distal metastasis, regional metastasis, locoregional metastasis, local spread, loc spread, locally spread, disseminated disease, disseminated ca, disseminated carcinoma, advanced disease, adv dz, adv ca, widespread disease, widespread mets, diffuse mets, systemic disease, systemic spread, systemic involvement, recurrent metastatic disease, recurrent mets, oligometastatic, oligo mets, oligo-metastasis, micrometastases, micro-metastases, micro mets,, macrometastases, macro-metastases, macro mets<br><br>evidence of mets, e/o mets, eo mets |

| | |
|---|---|
| | progression to metastatic, progression to mets, metastatic progression, metastatic spread, spread of mets, secondary tumor, secondary lesion, secondary ca, metastatic lesion, metastatic lesions, met lesion, met lesions, mets lesion, mets lesions<br>metastatic deposit, metastatic deposits<br>metastatic burden, tumor burden, metastatic involvement, involvement with mets, metastatic recurrence, recurrent mets<br><br>stage IV, stage 4, stg IV, stg 4, advanced stage, adv stage, adv stg, disseminated carcinoma, diss carcinoma, carcinoma with distant spread, ca with distant spread, extramedullary disease, extramed mets, EMD, widespread involvement, diffuse involvement |
| Custom Words | high grade, mass, malignant mass, metastatic mass, masses, progression, spread, masslike, lesion, malignancy, malignancies, carcinomatosis, tumor, cancer, neoplasm, nodule, effusion, tumors,<br>nodules, abnormal |

### Table S3. Keywords Used for Feature Extraction During GatorTron Fine-tuning

| | |
|---|---|
| Overall Metastasis Fine-Tuning | metastasis, metasasis, metastisis, metatasis<br>metastases, metasases, metatases<br>metastatic, metastitic, metastic, metatatic<br>metastasize, metastatize, metastisize, metatasize<br>metastasized, metastised, metastatized, metatasized<br>metastasizing, metastisizing, metatasizing<br><br>mets, mtas, msts, met's<br>met dz, met dis, met disease<br>met ca, m. ca |

| | |
|---|---|
| | adv dz, adv dis, adv disease, adv ca, adv cancer<br><br>bone metastasis, bony metastasis, bone mets, bony mets<br>osseous metastasis, oss mets, osseus metastases<br>lung metastasis, pulmonary metastasis, lung mets, pulm mets, pulmonary mets<br>liver metastasis, hepatic metastasis, liver mets, hepatic mets, hep mets<br>brain metastasis, cerebral metastasis, brain mets, CNS mets, cereb mets, intracranial mets<br>leptomeningeal metastasis, meningeal metastasis, leptomeningeal mets, meningeal mets<br>lymph node metastasis, nodal metastasis, LN mets, lymphatic mets, LN involvement<br>adrenal metastasis, adrenal mets<br>peritoneal metastasis, peritoneal mets, carcinomatosis, peritoneal carcinomatosis<br>pleural metastasis, pleural mets, pleural involvement, pleural spread<br>cutaneous metastasis, skin metastasis, cutaneous mets, skin mets<br><br>distant metastasis, distal metastasis<br>regional metastasis, locoregional metastasis<br>local spread, loc spread, locally spread<br>disseminated disease, disseminated ca, disseminated carcinoma<br>advanced disease, adv dz, adv ca<br>widespread disease, widespread mets, diffuse mets<br>recurrent metastatic disease, recurrent mets<br>oligometastatic, oligo mets, oligo-metastasis<br>micrometastases, micro-metastases, micro mets<br>macrometastases, macro-metastases, macro mets |

| | |
|---|---|
| | evidence of mets, e/o mets, eo mets<br>progression to metastatic, progression to mets, metastatic progression<br>metastatic spread, spread of mets<br>metastatic lesion, metastatic lesions, met lesion, met lesions, mets lesion, mets lesions<br>metastatic deposit, metastatic deposits<br>metastatic burden, metastatic involvement, involvement with mets<br>metastatic recurrence, recurrent mets<br><br>stage IV, stage 4, stg IV, stg 4<br>advanced stage, adv stage, adv stg<br>disseminated carcinoma, diss carcinoma<br>carcinoma with distant spread, ca with distant spread<br>widespread involvement, diffuse involvement |
| Brain/CNS Metastasis Fine-Tuning | brain metastasis, brain metastases, cerebral metastasis, brain mets, CNS mets, cereb mets, intracranial mets<br>mets to brain, mets to the brain, spread to brain, spread to the brain, brain involvement, brain met,<br>mets to cerebellum, cerebral involvement, spread to cerebellum, cerebral mets, cerebral met, cereb met<br>metastatic spread to brain, metastatic spread to the brain<br><br>leptomeningeal metastasis, meningeal metastasis, leptomeningeal mets, meningeal mets<br>leptomeningeal involvement, meningeal involvement, leptomeningeal spread, meningeal spread<br>leptomeningeal carcinomatosis, meningeal carcinomatosis, leptomeningeal met |

| | |
|---|---|
| | frontal lobe metastasis, frontal lobe mets, mets to frontal lobe, mets to the frontal lobe, frontal lobe met<br>spread to frontal lobe, spread to the frontal lobe, frontal lobe involvement, frontal lobe metastases<br><br>parietal lobe metastasis, parietal lobe mets, mets to parietal lobe, mets to the parietal lobe, parietal lobe met<br>spread to parietal lobe, spread to the parietal lobe, parietal lobe involvement, parietal lobe metastases<br><br>temporal lobe metastasis, temporal lobe mets, mets to temporal lobe, mets to the temporal lobe<br>spread to temporal lobe, spread to the temporal lobe, temporal lobe involvement, temporal lobe met<br><br>occipital lobe metastasis, occipital lobe mets, mets to occipital lobe, mets to the occipital lobe<br>spread to occipital lobe, spread to the occipital lobe, occipital lobe involvement, occipital lobe metastases<br><br>oligometastatic, oligo mets, oligo-metastasis<br>micrometastases, micro-metastases, micro mets<br>macrometastases, macro-metastases, macro mets<br><br>pachymeningeal carcinomatosis, dural metastasis, dural mets, mets to dural region, mets to the dural region<br>spread to dural region, spread to the dural region, dural involvement, dural metastases |

| | |
|---|---|
| | mets to dura, mets to the dura, spread to dura, spread to the dura, pachymeningeal metastasis pachymeningeal metastases, pachymeningeal met, pachymeningeal mets<br><br>spinal cord metastasis, spinal cord mets, mets to spinal cord, mets to the spinal cord spread to spinal cord, spread to the spinal cord, spinal cord involvement, spinal cord metastases<br><br>malignant brain lesion, intraparenchymal metastasis, intraparenchymal mets, intraparenchymal met intraparenchymal involvement, intraparenchymal metastases, brain secondary, brain secondaries<br><br>intracranial metastasis, intracranial mets, intracranial involvement, intracranial met, progression to brain,<br>lm mets, lm spread, spread to lm, lm carcinomatosis, lm involvement<br><br>subarachnoid metastasis, subarachnoid mets, mets to subarachnoid, mets to the subarachnoid, subarachnoid met<br>spread to subarachnoid, spread to the subarachnoid, subarachnoid involvement, subarachnoid metastases<br><br>intradural metastasis, intradural mets, mets to intradural region, mets to the intradural region spread to intradural region, spread to the intradural region, intradural involvement, intradural metastases |

### Supplemental Material 1. Prompt Template used for Prediction of Overall Metastasis

Determine whether the clinical note states that the patient has metastatic cancer.

Do not infer potential metastasis. Strictly use the context provided in the note.

Answer only in one line:

Metastasis: YES

or

Metastasis: NO

Do not explain further.

Clinical Note:

{note}

Answer:

### Supplemental Material 2. Prompt Template used for Prediction of Brain/CNS Metastasis

"Brain" was replaced with "CNS" depending on reference standard labels.

Determine whether the clinical note states that the patient has brain metastasis.

Ignore other sites of metastasis. Strictly use the context provided in the note.

Answer only in one line:

Brain metastasis: YES

or

Brain metastasis: NO

Do not explain further.

Clinical Note:

{note}

Answer:

### Table S4. GatorTron Error Analysis for Overall Metastasis Prediction

| Error Category | Frequency (%) | Justification | Comparison to MedGemma & Llama3 |
|---|---|---|---|
| Ambiguous Text | 10 (33.3%)) | Four false positive and six false negative cases had ambiguous phrasing, including “concern for, “possible,” and “suspicious” metastatic disease without confirmation of positive or negative metastasis presence. | Two false positive predictions aligned with Llama3. One false positive prediction aligned with MedGemma. All other predictions by Llama3 and MedGemma aligned with registry labels. |
| Correct Prediction | 4 (13.3%) | All false positive cases had clear mentions of metastasis presence. Ex: “Recent diagnosis of right lung adenocarcinoma with lymph node metastasis.” | All predictions aligned with Llama3. One positive prediction was missed by MedGemma but predicted by GatorTron and Llama3. |
| Missing Text | 3 (10.0%) | All false negative cases had no mention of metastasis presence, indicating the possible presence of additional, unavailable notes. | MedGemma’s and Llama3’s predictions aligned with GatorTron’s |
| Incorrect Prediction | 13 (43.3%) | One false positive case had a clear mention of “no adenopathy or metastatic disease.” Two false negative cases had clear mentions of “malignant effusion” and “lung cancer with mets,” respectively. | Three false positive predictions aligned with Llama. All predictions by MedGemma aligned with registry labels. |

### Table S5. MedGemma Error Analysis for Overall Metastasis Prediction

| Error Category | Frequency (%) | Justification | Comparison to Llama3 & GatorTron |
|---|---|---|---|
| Correct Prediction | 9 (39.1%) | All false positive cases had clear mentions of metastasis presence. | Llama3 identified all but two cases as positive. GatorTron identified four cases as positive. |
| Missing Text | 6 (26.1%) | All false negative cases had no mention of metastasis presence, indicating the possible presence of additional, unavailable notes | Llama3 identified all cases as negative. GatorTron identified only half of the cases as negative, despite no indication of metastasis presence in the notes. |
| Ambiguous Text | 6 (26.1%) | There were four false positive and two false negative predictions. In all cases there was mention of "probable" metastasis or cases in which a lesion "may represent" malignant spread without official diagnosis of metastasis. | Llama3 produced the same prediction for three false positive predictions, while GatorTron produced the same prediction for one false negative prediction. |
| Incorrect Prediction | 2 (8.7%) | One false positive prediction was produced, despite clear indication of diagnoses metastasis. One false positive prediction was produced, despite no evidence of metastasis following imaging. | Both cases were predicted correctly by Llama3 and GatorTron |

### Table S6. GatorTron Error Analysis for Brain/CNS Metastasis Prediction

| Error Category | Frequency (%) | Justification | Comparison to GatorTron & MedGemma |
|---|---|---|---|
| Missing Text | 4 (28.5%) | All false negative cases had no mention of metastasis presence or no mention of the brain or CNS regions, indicating the possible presence of additional, unavailable notes. | All of Llama3 and MedGemma predictions aligned with GatorTron's. |
| Ambiguous Text | 1 (7.1%) | One false positive case, in which the documentation indicated "suspected" metastasis to the brain. | Llama3 and MedGemma aligned with the registry label. |
| Incorrect Prediction | 10 (71.4%) | Five false positive cases either mentioned metastasis to other sites or did not indicate metastasis at all. Five false negative cases demonstrated clear presence of metastasis in the text. | All predictions by Llama3 and MedGemma aligned with registry labels. |

### Table S7. Llama3 Error Analysis for Brain/CNS Metastasis Prediction

| Error Category | Frequency (%) | Justification | Comparison to MedGemma & GatorTron |
|---|---|---|---|
| Missing Text | 4 (80%) | All false negative cases had no mention of metastasis presence or no mention of the brain or CNS regions, indicating the possible presence of additional, unavailable notes. | All of Llama3's predictions aligned with MedGemma's. One prediction by GatorTron aligned with the registry label, despite no indication of metastasis in the note. |
| Incorrect Prediction | 1 (20%) | One false positive prediction was produced despite no documented presence of metastasis to the brain/CNS regions. | GatorTron and MedGemma predicted this case accurately. |
